# The Impact of Martial Arts on Mental Health

**DOI:** 10.1101/2023.09.17.23295679

**Authors:** Rick A. Hiciano, Brian J. Piper

## Abstract

Sports Psychiatry has been developing for the last three decades and is growing to be an integral part of Sports Medicine and Psychiatry. With the rising growth of mental health awareness in popular culture, and public announcements of professional athletes describing their struggles with mental health, the need for Sports Psychiatrists is becoming more apparent. However, limited data on mental health disorders amongst athletes poses issues in terms of providing standardized and reliable interventions for treatment. This is especially the case in less researched sports, such as martial arts. The current study aims to investigate the relationship between mental health and martial artists, specifically evaluating the relationship between domains of mental health, including aggression, depression, anxiety, and self-esteem, and Jeet Kune Do/mixed martial arts practitioners. The study will take place at Garcia Gung Fu, a martial arts dojo in Harlem, New York in addition to recruits from a Reddit Martial Arts form. Participants will include both adults and children. The findings in this study will contribute to the body of literature in both sports medicine and psychiatry. Additionally, this study will further illustrate the importance of the field of sports psychiatry and well as highlight the need for improved interventions for athletes.

## Introduction

The Impact of Martial Arts on Mental Health

### Background

The field of Sports Psychiatry has recently emerged and has been developing for the last three decades and is growing to be an integral part of Sports Medicine^1^. This is widely due to the emerging recognition and awareness that athletes suffer from mental health issues and can benefit from treatment without hindering but improving their academic performance. Sports Psychiatry is a clinical practice aimed at treating athletes and the mental health disorders that affect them using a psychotherapeutic foundation integrated with medication. As recently published in the emerging field, Sports Psychiatrists are equipped to treat a range of mental health problems from mild to severe to mitigate the risk of those problems and potentially prevent career-ending illness if not properly managed^2^. A Sports Psychiatrist - the term coined by J.H. Massamino in 1987 - can provide an extensive range of services to the athletic community. In fact, it is become more prevalent that Sport psychiatrists occupy significant advisory positions within organizations governing Olympic sports, such as the International Federation of Aquatic Sports; collegiate sports, such as the National Collegiate Athletic Association; professional sports organizations, such as the National Football League and the PGA (Professional Golfer’s Association)^3^. Similarly, modern day NBA players such as Demar DeRozan, Kevin Love, and Metta World Peace have even spoken publicly about their struggles with mental health, with Metta World Peace thanking his psychiatrist after his earning his 2010 NBA championship and Kevin Love providing an exclusive, hour-long public interview about his battle with depression in 2018. With mental health awareness on the rise both in the general public and among elite athletes, the need for Sports Psychiatrists is only growing.

As stated previously, mental health risks, including depression and anxiety, are widely known to affect athletes. One meta-analysis concluded that anxiety and depression was prevalent amongst 34% of former and current elite athletes of both American and Olympic sports^4^. Another study concluded that anxiety symptoms and other determinants of mental health found in athletes were comparable to that of the general population^5^. This study strengthens the claims that participation in sports, including but not limited to soccer, American football, basketball, baseball, handball, volleyball and gymnastics, even at an elite level, does not protect individuals from health disorders^5^. Another meta-analysis concluded that elite level athletes are vulnerable to a range of mental health disorders related to both sporting and nonsporting factors^6^. This study compiled a search of over two thousand articles and settled on sixty high quality papers after their exclusion process. They synthesized the limited published reviews on the evidence and incidence of mental health concerns amongst athletes, though at the time there were no high-quality reviews on the psychological and mental wellbeing of elite level athletes^6^. Most studies utilized in this meta-analysis included self-report data of elite level athletes, which leaves the prevalence of diagnosed mental health disorders amongst both elite and amateur athletes in question. This study seems to be in stark contrast with the popular belief that sports exert positive physical, and often social, benefits that protects individuals from mental health concerns. Similarly, an executive summary suggests that athletes face unique risk factors for mental health disorders in comparison to the general population^7^. Furthermore, given the limited data on worldwide, popular and televised sports, it is less likely that there will be current literature on less well-known sports or their variations including forms of martial arts (i.e. Jeet Kune Do, a form of martial arts). These studies make it suffice to say that participation in sports often overshadows concerns for any individual mental health burden, due to widespread belief of the positive associations attributed to sports. It is no question that sports promote healthy, positive behaviors, but it is not entirely known exactly in which type of individual these benefits can be seen or expected. Furthermore, the question of why some athletes experiences an exacerbation of mental health concerns in sports known to promote positive characteristics is raised. Regardless, it is the popular belief that sports inherently promotes positive mental health is what led to the challenge to the idea that being an elite athlete, or even a highly skilled amateur, will protect one from the emotional and mental strain^8^. This study concluded that the current evidence demonstrating the prevalence of mental health disorders amongst elite level athletes is enough to consider revision of the current mental health prevention and treatment strategies in place for athletes^8^.

Like other sports, there is a widespread belief that participation in martial arts, a combat sport, has specifically illustrated many positive outcomes for its practitioners. Another meta-analysis demonstrated that there is evidence for martial arts as a mental health intervention that can be used for improving well-being and reducing symptoms of mental health disorders^9,10^. As cited, some of these improvements include a reduction of aggression in males, a decrease in the prevalence of anxiety and depression, and an increase in self-esteem amongst practitioners^9, 10,11^. Specifically, these improvements were seen in practitioners of tae kwon do, karate, and tai chi^11, 12, 13, 14, 15, 16^. With martial arts being a combat sport, it might be surprising that a sport centered in willful violence can serve as a protective factor for mental health, while other sports questionably display this same trend. In fact, several studies show contradicting results including seeing no significant difference seen in self-esteem improvement amongst tae kwon do practitioners of various experience levels^17^. Another study showed an increase rather than an expected decrease in levels of aggression among martials arts practitioners^18^. With these contradictory findings, limited research, and possible selective interpretations of earlier studies, it is difficult to determine the validity of the current data on the impact martial arts has on mental health^9,10^. For that reason, this study serves to broaden the current literature and introduce both jeet kune do and mixed martial arts to the developing field of sports psychiatry. This study can serve to confirm the findings of previous studies and potentially elaborate on the relation between martial arts and mental health.

Specifically, this study will investigate and analyze the impact that Garcia Gung Fu, an urban, inner-city Jeet Kune Do/Mixed Martial Arts dojo has had on the mental health of its students. Garcia Gung Fu is a martial arts school located in Harlem, New York City. Harlem is known for being an area full of different cultures made up of immigrants and local residents. Harlem is an impoverished area with 28% of its residents, more than double the rate in the US, falling under the US poverty line^19^. The instructor, known as Sifu Garcia, instructs Jeet Kune Do, mixed martial arts and self-defense to not only train fighters for competition, but to provide self-defense training for his students who face the daily stressors of living in a low-income community with a high rate of violence. Jeet Kune Do is the art of street self-defense in combination with techniques from various styles and disciplines of martial arts. Sifu Garcia synthesizes many styles of martial arts including but not limited to jujitsu, kick boxing, muy thai, wrestling, street fighting and self-defense. The synthesis of these disciplines makes up what is known as Jeet Kune Do, coined and created by Bruce Lee, and modern day mixed martial arts. From the year 2020 - 2021, there was an 11% increase in violent crimes, with an overall crime rate of 762 violent crimes committed per 100,000 people in New York County in the year 2021^20^. This does not include other crimes such as crimes with firearms, index crimes, and property crimes. With these rates of violence, Sifu Garcia provides invaluable martial arts teachings that not only provide a safe haven for his students but provides them with the tools to feel safer in their environment. He not only teaches physical survival techniques, but mental and spiritual techniques as well.

Given the high rate of violence and poverty within New York City, it is no question that there are several risk factors predisposing adults and children to various mental health disorders. There is widespread evidence in the literature showing that increased exposure to violence in disadvantaged communities increases levels of depressive symptoms in individuals, particularly adolescents. Furthermore, it’s been shown that increased exposure to violence in the family increased depressive symptoms during the high school years of a person’s adolescence^21^. Similarly, exposure to violence predisposes individuals to mental health concerns, especially youth coming from lower socioeconomic status^22^. With Garcia Gung Fu as the focus, this study will analyze how the practice of Jeet Kune Do/Mixed Martial arts has influenced the prevalence of mental health disorders among its students. Specifically, the aim is to detect whether training this art will influence the prevalence and severity of anxiety, depression, violence, aggression, self-esteem and safety perception levels among both adult and pediatric practitioners. Given that martial arts is a combat sport, it will be interesting to determine how perceived violence in martial arts relates to mental health.

### Key Word Definitions

#### Martial Arts

According to the Encyclopedia Britannica, *martial arts* is defined as a system of combat that can be practiced for various reasons including self-defense, military or law enforcement, competition, entertainment or physical, mental and spiritual development. For this study, we will adhere to this definition as we intend to include practitioners of martial arts regardless of their reason for practice.

#### Aggression

Aggression is a subjective feeling that may take various forms. For that reason, we will define aggression according to Oxford’s Learner’s dictionary: aggression is a feeling of anger or hate that may result in violent or threatening behavior.

#### Violence

When the word violence arises, the common idea is to think of a fight, physical abuse, or anything that involves the physical or emotional dominance of another living being. Since violence can entail various domains, we will use the term violence as defined by the Oxford Learner’s dictionary: violence is the exercise of physical and/or emotional force and/or energy used to inflict injury on, or cause damage to, persons or property.

#### Self-esteem

In this study, self-esteem will be defined according to the Oxford Learner’s dictionary, so as not to confuse it with self-confidence or another measure of self-perception: self-esteem is defined as a feeling of being happy with your own character and abilities.

### Study Aims

This study aims to contribute and expand on the current literature within sports medicine and psychiatry. This is a novel study that will not only contradict or confirm existing literature, but potentially establish new relationships between various domains of mental health and the practice of martial arts. The specific objectives are listed below:

1. To determine the relationship between martial arts training and individual perception of safety within an urban environment.
2. To determine and clarify the relationship between martial arts training and the future development of depression and anxiety.
3. Establish whether there is an increased or decreased level of aggression in martial artists of urban environments in comparison to the general population.
4. Analyze the correlation between Adverse Childhood Experiences with the development of aggression, depression and anxiety within martial artists
5. Evaluate the levels of self-esteem of martial arts students.

## Methods

### Selection Criteria

References were obtained from various journals including the American Psychological Association, The American College of Sports Medicine, the British Journal of Sports Medicine, and the Journal of the American Medical Association. Articles and journal entries were searched from various databases such as PubMed, PsychINFO, ScienceDirect, and the National Institute of Health. Search terms included “Mental Health,” “Martial Arts,” “Sports Psychiatry,” “Jeet Kune Do,” “Aggression,” “Childhood Adverse Experiences,” and “Safety Perception.” Publication years included 1960 – 2022. Only peer-reviewed articles were selected. As articles of interest were found, more related articles were searched for and chosen from their references section.

### Institutional Review Board (IRB)

Currently, this study is pending IRB approval by Geisinger Institutional Review Board. The study number is: 2023-1055.

### Participants

Participants chosen for this study will include both adults and children currently and formerly training at Garcia Gung Fu, and individuals from the general population. We aim to have at least 30 participants making up the experimental group and at least 30 participants in the control group in our study. These will include individuals aged seven and over. There will be two experimental groups, one composed of children and one of adults.

There will also be two control groups, one composed of children and one of adults. Participants making up the control group will be individuals living in the United States who may or may not already participate in martial arts. These individuals will be recruited from local universities, the local community, family and friends of the participants and a reddit martial arts forum. All individuals not currently training at Garcia Gung Fu, but who are found to currently practice martial arts at a different site not affiliated with Garcia Gung Fu will make up another group that can be used as another comparison group but not as the control. Only those individuals who do not train martial arts or who have trained martial arts for a total of less than 3 months will serve as the control.

All participants are expected to have a basic 6-8^th^ grade reading level and a basic understanding of the English language. For minors or other individuals below the standard age for a 6^th^ grade reading level (average age of 11 years or under as determined by US Department of Education), a research investigator will assist in reading and responding to the required assessments if there is confusion or difficulty on behalf of the participant. Similarly, since individuals will be recruited from an economically disadvantaged area, it is a possibility that some individuals may have a reading level below 6^th^ grade. In this instance, a research investigator will assist the participant in reading through the required materials. The required assessments have easy readability and comprehension and therefore there is no expectation that most participants will struggle in completing them. Any form of coercion or undue influence will be minimized by explaining the purpose of the study, answering any questions, emphasizing that participation is voluntary and will not be used for any form of diagnosis or medical reporting, and that all data will be kept anonymous. Participants will also be reassured that none of the information obtained will be submitted to any health or employment institutions and that choosing to participate will not affect their health or employment standing in any way.

#### Protocol 1

##### Recruitment

###### Experimental Group

Participants for the experimental groups will be approached in-person at Garcia Gung Fu and will be asked to participate in the study. Previous students and other students connected to Garcia Gung Fu from other martial arts institutions may also be recruited in-person and be included as part of the experimental group. All adults and parents of children will be asked directly for their participation and will be briefed as to the nature of the study, specifying that this is a study analyzing mental health trends among martial artists. If permission is granted, all participants will be provided with the option of a paper or electronic version of the questionnaires to be completed. Upon recruitment, all participants will be told in the briefing that there is no compensation for their participation, that their participation is voluntary, that they can quit at any point, and that their participation will contribute to the potential development in the field of medicine.

###### Control Group

Individuals making up the control group will be recruited directly in-person. Individuals approached directly will include the family and friends of individuals training at Garcia Gung Fu. Those individuals will be provided with the option of an electronic or paper version of the questionnaires to be completed. Upon recruitment, all participants will be told in the briefing that there is no compensation for their participation, that their participation is voluntary, that they can quit at any point, and that their participation will contribute to the potential development in the field of medicine.

##### Inclusion criteria

###### Experimental group

All individuals aged 7 and over that currently trains at Garcia Gung Fu or a connected martial arts institution for at least 3 months

###### Control group

Any individual aged 7 and over that has never trained martial arts or have trained for less than 3 months.

##### Exclusion criteria

###### Experimental Group

Individuals that have trained martial arts for a total of less than 3 months or are under the age of 7.

###### Control Group

Individuals that have trained martial arts for a total of over 3 months and are under the age of 7.

##### Procedure for protocol 1

###### Experimental Group

There will be two experimental groups for individuals recruited from Garcia Gung Fu, one consisting of children and the other of adults. For in-person administration, participants will first be asked for permission and be required to sign a consent form prior to the administration of the surveys. A research assistant will go over the consent form with the potential participant. Once consent is obtained, participants will be briefed on the nature of the study and be reassured that all of the information will be de-identified and used for research purposes only. No data will be used to assess or diagnose any participant for any disorder. No data will be used for reporting to legal authorities. Once understood, a research associate will provide the surveys to the participant and be within reach to answer any questions that arise. For individuals who opt to complete the study questionnaires online, they will first be required to sign a consent form with the research associate. Once consent is obtained, individuals will be provided with the electronic version of the questionnaires. Completion of the surveys and demographic information should estimate an hour in length. If any child or adult has difficulty understanding any of the required survey materials, a research associate will be available to provide clarification. All surveys will include an ID number consisting of a randomly generated 6-digit number. Once complete, the research associate will collect the survey and store them in a secure folder. Once complete, google will store the survey, locked by a pin.

###### Control Group

There will be two control groups, one consisting of adults and the other of children. The control groups will consist of individuals recruited in person and the same procedure used for the experimental group will be used for this control group.

#### Protocol 2

##### Recruitment

###### Experimental Group

Additional participants making up the experimental group consisting of adults will include interested individuals responding to a post in the martial arts group section of Reddit. The post will briefly describe the nature of the study, its purpose, a link to a waiver of consent written as a statement that allows the participant to choose to “agree or not agree” to waive their consent, and the survey/questionnaire materials will be included in the post. All individuals acquired from reddit will be aged 18 and older, in accordance with reddit guidelines and will need to agree to being aged 18 or over when submitting the study materials. Participants will be sorted based on the inclusion/exclusion criteria stated. Children or vulnerable populations will not be used for this experimental group. Upon recruitment, all participants will be told in the briefing that there is no compensation for their participation, that their participation is voluntary, that they can quit at any point, and that their participation will contribute to the potential development in the field of medicine.

###### Control Group

Additional Individuals making up the control group consisting of adults will be recruited via the reddit martial arts forum and will be sent an electronic version of the required materials in addition to a waiver of consent to maintain anonymity. All individuals will be briefed as to the nature and purpose of the study, specifying that this is a study analyzing mental health trends among martial artists. A link to a waiver of consent written as a statement that allows the participant to choose to “agree or not agree” to waive their consent and the survey/questionnaire materials will be included in the post. All individuals acquired from reddit will be aged 18 and older, in accordance with reddit guidelines. Participants will be sorted based on the inclusion/exclusion criteria stated. Children or vulnerable populations will not be used for this control group. Upon recruitment, all participants will be told in the briefing that there is no compensation for their participation, that their participation is voluntary, that they can quit at any point, and that their participation will contribute to the potential development in the field of medicine.

##### Inclusion criteria

###### Experimental group

All individuals aged 18 and over that currently and/or have trained martial arts of any discipline for at least 3 months.

###### Control group

All individuals aged 18 and over that have never trained martial arts and/or have trained martial arts of any discipline for less than 3 months.

##### Exclusion criteria

###### Experimental Group

Individuals that have trained martial arts for a total of less than 3 months and/or are under the age of 18.

###### Control Group

Individuals that have trained martial arts for a total of over 3 months and/or are under the age of 18.

##### Procedure, protocol 2

For individuals recruited by Reddit, participants will click the link located in the post describing the study, which will send them to a google form containing a waiver of consent and the survey/questionnaires required. The waiver of consent will be required for submission of the study materials. No identifiable information such as email, name or signature will be asked or obtained. Individuals will be sorted based off the inclusion/exclusion criteria described. Once complete, google will store the survey, locked by a pin.

##### Data Storage

All data will be stored in a secure, locked google document. Participant names will not be included with any of the information. All physical data will be transcribed to an online source, with the physical data being shredded and discarded. All participants will be assigned a randomly generated number associated with their completed questionnaire answers. In the event a participant wishes to withdraw from the study before or after the completion of the required materials, that information will be discarded and will not contribute to the results. Physical information will be kept in a locked area. Electronic data will be kept in a locked google folder. Only IRB-study approved staff will have access to this information. Records of data obtained will be de-identified and kept indefinitely and can be used for other IRB approved research.

##### Study Duration

Participant’s involvement in the study will take approximately an hour in length or as long as needed to complete the required study questionnaires but not to exceed two hours. The entire study will run for approximately 3 months.

##### Assessments

Multiple assessments will be used to measure various subsets of mental health within participants. These are listed below:

###### 1) The Buss-Perry Aggression Questionnaire^23, 24^

The Buss-Perry Aggression Questionnaire (BPAQ) consists of 29 self-administered items rated on a 5-point Likert scale. The BPAQ includes four subscales: Physical Aggression (items 1-9), Verbal Aggression (items 10-14), Anger (items 15-21), and Hostility (items 22-29.) This Questionnaire has become the gold standard in measuring aggression, and its validity and reliability has been tested in various populations.

###### 2) Patient Health Questionnaire - 9 (PHQ-9)^25, 26^

The PHQ-9 is a self-administered, 9 item depression module that detects depression and measures its severity. It has been widely used in clinical settings for depression screening and has been widely tested for reliability and validity in the literature.

###### 3) Generalized Anxiety Disorder Screener −7 (GAD-7)^27^

The GAD-7 is a self-administered, 7-item anxiety questionnaire that detects anxiety and measures its severity. It has been widely used in clinical settings for Generalized Anxiety screening and has been widely tested for reliability and validity in the literature.

###### 4) Rosenberg Self-Esteem Scale^28^

This is a 10-item scale that measures global self-worth by measuring both positive and negative feelings about the self. It features a 4-point Likert scale ranging from strongly agree to strongly disagree. This scale has been widely used as a self-report measure of self-esteem since its publication in 1962 and is both reliable and valid.

###### 5) Child Rosenberg Self-Esteem Scale^29^

Like the original Rosenberg Self-Esteem Scale, the Child Rosenberg Self-Esteem Scale (CRSES) has been adapted to include simplified language to adapt to the understanding of children aged 12 years and younger. This scale includes a 10-item list that can be measured on a 4-point Likert scale ranging from “Very True” to “Definitely Not True.” This has been tested for and found to be a reliable and valid tool.

###### 6) Perception of Safety Tool from Rijswijk and Haans adapted from Haans & de Kort (2012)^30, 31^

This Safety Perception tool is an adaptation of a tool used in the field of Environmental Psychology to determine an individual’s self-perceived level of safety within one’s environment. This is a 3-item tool measured from 1 (Very unsafe) to 5 (very safe).

###### 7) Adverse Childhood Experiences Questionnaire (ACEs)^32, 33, 34^

The ACEs Questionnaire has been widely used in the literature and in clinical practice for prediction of physical and mental health disorders and for informing possible treatment interventions in adults and children. This is a valid and reliable tool that can help determine the number of risk factors predicting the development of mental health disorders. This questionnaire will only be administered to adults aged 18+ in the study. The authors of the study are mandated reporters, and because deidentified data is being used, there is no way to report suspected abuse in children without breaking confidentiality. Likewise, we want to minimize cultural insensitivity that may arise due to differences in views on discipline.

###### 8) Demographics

Demographics to be collected will include age, ethnicity/race, sex/gender, length of time training martial arts (both at Garcia Gung Fu and other institutions) and household income (to determine socioeconomic status).

## Results

For analysis of data, inferential statistics, including independent samples T-test or chi-squared tests, will be run to analyze for any differences in levels of aggression, depression, anxiety, self-esteem and perception of safety between martial arts practitioners vs. non-practitioners. Given the number of questionnaires being used, there will be an opportunity for a wide range of data from which to extrapolate.

A Pearson’s Correlational analysis may be used to draw a relationship between the length of training, childhood adverse experiences and severity of any of the subsets/measures for mental health.

## Discussion

With the rise of sports psychiatry, there is room for a deeper scientific and psychological investigation of the ailments and diseases that target many athletes across a variety of sports. Given the incongruency between popular belief of sports as a protective factor for mental health, and sports as a potential exacerbation of mental health, this study will help deconstruct any misconceptions, confirm previous literature, and potentially illuminate new relationships that can suggest possible clinical interventions. Specifically, the sport of martial arts will be more thoroughly investigated. This is important as mental health and combat sports are trending topics in popular culture. Additionally, more professional athletes across different sports, including martial arts, are speaking out publicly on mental health and thus raising awareness of the need for more mental health assistance amongst athletes. This increasing need, along with the potential findings of this study, will assist in strengthening the role of Sports Psychiatry in the life of an athlete.

### Potential Limitations

Since the data required in this study will be self-reported, there is the possibility of self-report biases including recall bias. As participants will be going off their own understanding, which may vary depending one an individual’s age, education and general background. Factors such as age, socioeconomic status and education level will try to be controlled, but there is no controlling for the subjective feelings that may be asked in the questionnaires.

Since the questionnaires do ask questions related to the subjective feelings and experiences of the participant, there is the possibility of social desirability bias occurring. De-identification of participant information and reassurance of confidentiality will be used to control for this bias.

Participants partaking in martial arts may already have a predisposition towards better mental health, as the nature of martial arts draws individuals towards believing in its ancient principles. Similarly, participants partaking in martial arts may have the means to afford the classes, potentially placing them above the average socioeconomic status of local residents. This can be a confounding variable when comparing the experimental group to the control.

Given the size of the martial arts site, there may be a limited sample size depending on willing participation. Attempts to recruit participants from other associated martial arts schools will be used as a method to control for this.

Since martial arts has various styles and philosophies, controlling for standardized instruction poses an issue of external validity. Future interventions will be needed to provide a standardized model that can control for differences in teaching abilities and experience. Currently, there is no known method for standardizing instructor training across martial arts. This study will illuminate the need for this as a potential therapy model.

## Data Availability

All data produced in the present study will be made upon reasonable request to the authors.

## Acknowledgements.

I would like to acknowledge Dr. Brian Piper for his ongoing guidance and support as my faculty mentor.

## Disclosures

None.

## Notes

### Competing Interest Statement

The authors have declared no competing interest.

### Funding Statement

This study did not receive any funding.

### Author Declarations

Geisinger Institutional Review Board of Geisinger Commonwealth School of Medicine gave ethical approval for this work.

